# Calcium-sensing receptor antagonism as a novel therapeutic for pulmonary fibrosis

**DOI:** 10.1101/2020.03.12.20034751

**Authors:** Kasope L. Wolffs, Bethan Mansfield, Richard T. Bruce, Ping Huang, Martin Schepelmann, Sarah C Brennan, Line Verckist, Dirk Adriaensen, Rachel Paes De Araújo, Luis A.J. Mur, Richard Attanoos, Keir E. Lewis, Jeremy P.T. Ward, Christopher J. Corrigan, Paul J. Kemp, Benjamin D. Hope-Gill, Daniela Riccardi

## Abstract

**Background:** Idiopathic pulmonary fibrosis (IPF) is a disease with very poor prognosis and no curative therapies. The G protein-coupled, calcium/cation-sensing receptor (CaSR) is activated by environmental pollutants and by arginine-derived polyamines, which are thought to play a role in IPF. Whether the CaSR is involved in the pathogenesis of pulmonary fibrosis is unknown.

**Objective:** To investigate the CaSR as a novel drug target for the treatment of pulmonary fibrosis (PF).

**Methods and results:** CaSR protein expression is found in the airway epithelium in the neuroepithelial bodies of the healthy and IPF human lung. Expression of arginine pathway-linked polyamines is increased in PF patient saliva samples compared to non-PF patients. Arginine pathway metabolites, ornithine and spermine, activate the CaSR in primary normal human lung fibroblasts (NHLF), effects prevented by CaSR antagonism using the calcilytic NPS2143. In NHLF calcilytic also reversed the pro-fibrotic effects of exogenous TGFβ1 administration on Rho kinase and αSMA expression, proliferation, collagen production and IL-8 secretion. Targeted CaSR ablation from fibroblasts and smooth muscle cells protects mice from spontaneously occurring, age-related lung fibrosis.

**Conclusions:** Sustained CaSR activation in the lung drives pro-fibrotic processes, which can be reversed by calcilytic. Pharmacological and genetic CaSR blockade reduce both TGFβ1-induced and naturally occurring pro-fibrotic changes. This work provides the scientific rationale for developing inhaled calcilytics as novel therapeutics for IPF.

**KEY MESSAGES:** *Key question:* How does the calcium/cation-sensing receptor (CaSR) promote pulmonary fibrosis?

*Bottom line:* The CaSR is expressed in human IPF and experimental models of PF where receptor inhibition prevents pro-fibrotic changes and pulmonary remodeling.

*Why read on:* CaSR blockers, calcilytics, represent a novel treatment for IPF.

## INTRODUCTION

Pulmonary fibrosis occurs in the setting of various interstitial lung diseases of known but mostly unknown (idiopathic) etiologies. The most common type, idiopathic pulmonary fibrosis (IPF) has a worse 5-year survival rate (25%) than many cancers.[1] There is a pan-European increase in the incidence of IPF, with the UK and Finland recording the greatest rise in mortality.[2] There is currently no cure for IPF, and lung transplantation still represents the best therapeutic option which restores lung function and improves survival. However, most patients are precluded from transplantation due to their advanced age/frailty and the presence of co-morbidities such as pulmonary arterial hypertension (PAH), emphysema and lung cancer.[3, 4] Presently, the only therapeutic options are anti-fibrotic agents, pirfenidone and nintedanib.[5] These agents can, in some patients, slow decline in lung function,[3] but have significant adverse effects and do not directly target IPF co-morbidities.[3, 4] Thus, there is an urgent need to develop new therapeutics with fewer side effects, which improve symptom management and increase patient survival.

The incidence of IPF increases with age.[6] In younger people, fibrosis is a beneficial self-resolving wound-healing response to organ injury.[7] However, as aging occurs, this process is hijacked, leading to deleterious effects in the lungs, heart, kidney and liver.[7, 8] Although the specific insults initiating the disease in different organs may vary, the path to organ fibrosis is similar, involving the activation of multiple cell types and a complex network of signaling cascades, many of which are associated with G protein-coupled receptors (GPCR)[9] located on a number of cell types. These cells include epithelial cells (thought to be the site of the initial injury), inflammatory cells, (myo)fibroblasts (the fibrogenic effectors), and pulmonary neuroepithelial cells, which release mitogenic factors that may act as a signal for epithelial and fibroblast proliferation.[8–10] Most of these cells produce TGFβ which mediates key fibrotic hallmarks such as aberrant fibroblast proliferation, myofibroblast accumulation and extracellular matrix (ECM) deposition.[11] One way that TGFβ could exert its pro-fibrotic effects is *via* the calcium/cation-sensing receptor (CaSR). The CaSR is a member of group C GPCR and it is expressed in the lung, where it acts as a multi-modal chemosensor for components of environmental pollutants (*e*.*g*. urban particulate matter, Ni^2+^ and Cd^2+^) and polycations (*e*.*g*., eosinophil cationic protein, major basic protein). CaSR is also activated by various polyamines (*e*.*g*. putrescine, spermine and spermidine), and basic amino acids (*e*.*g*. L-arginine and L-ornithine) which are particularly abundant in the lung.[12, 13] Expression of these activators increases in IPF patients.[14] CaSR activation leads to an increase in intracellular Ca^2+^ (via G_q/11_ coupling),[12] increase in Rho kinase-mediated actin stress fiber assembly (via G_q/11_ and possibly G_12/13_ coupling)[15] and to a decrease in intracellular cAMP levels (via G_i/o_ signaling).[16] Previously, CaSR signaling has been implicated in the remodeling and proliferative process of several diseases including asthma;[16] cardiac fibrosis;[17] pulmonary arterial hypertension (PAH);[18] and chronic obstructive pulmonary disease (COPD).[19] Pertinently, the last two diseases are common IPF comorbidities. Whether polyamines, acting at the CaSR, also play a role in IPF has never been studied.

This study aims to investigate CaSR expression in healthy and IPF lungs, the role of polyamines in IPF pathogenesis and the ability of CaSR inhibitors, calcilytics, to suppress TGFβ-induced pro-fibrotic effects in primary human lung fibroblasts. The involvement of CaSR in age-related pulmonary fibrosis is investigated in ageing mice with targeted CaSR ablation from SM22α-positive cells, leading to CaSR ablation from fibroblasts and smooth muscle cells.

## METHODS

### Human studies

#### CaSR expression in human control and IPF lung tissue

Immunohistochemistry was carried out as described previously[20] in 5 µm-thick paraffin paraformaldehyde-fixed biopsies from human lungs. Consecutive sections were incubated overnight with different primary antibodies (rabbit anti-human GRP polyclonal Ab, 1:1000, #ab22623; mouse anti-human CaSR mAb, 1:200, #ab19347; rabbit anti-human αSMA polyclonal antibody, 1:100, #ab5694; all Abcam, Cambridge, UK). The sections were incubated with the secondary antibodies (DAM-BIOT, 1:500; DAR-BIOT, 1:500; Jackson ImmunoResearch, Cambridgeshire, UK) followed by ExtrAvidin-horseradish peroxidase (HRP, 1:1000 in PBS; Sigma, Dorset, UK). HRP enzyme activity was visualized using 3,3’-diaminobenzidine (DAB; Dako, Heverlee, Belgium) as a chromogen. Cell nuclei were counterstained with haematoxylin. Negative controls were performed by preabsorption and replacement of the CaSR antibody with custom-made CaSR-ADD antigenic peptide and IgG2a isotype control, respectively.

#### Targeted metabolite fingerprinting by flow infusion electrospray high-resolution mass spectrometry (FIE-HRMS)

FIE-HRMS was used to evaluate the potential of saliva as a source of non-invasive polyamine biomarkers for PF. Spontaneous saliva was collected and processed from 6 patients with PF (mean age 81 ± 5 yrs, 67% male) and 6 controls without lung disease (mean age 70 ± 14 yrs, 33% male). PF diagnosis was confirmed at regional multidisciplinary team meetings. No statistical differences in age or gender were observed between groups.

Samples were processed and analyzed by FIE-HRMS as described previously.[21] FIE-HRMS was performed using a Q executive plus mass analyzer instrument with UHPLC system (Thermo Fisher Scientific, Bremen, Germany), where mass/charge (m/z) were generated in positive and negative ionization mode in a single run. All samples were run in duplicate with no significant differences between duplicates.

### *In vitro* studies in primary human lung fibroblast

#### Cell culture

Human primary lung fibroblasts from normal patients (NHLF, CC-2612; passage 2-4) were purchased from Lonza (Slough, UK) and cultured in fibroblast growth medium (FGM-2) containing (CC-3131). See online supplementary material.

#### Measurements of [Ca^2+^]

CaSR-induced changes in [Ca^2+^]_i_ were measured in fura-2 loaded individual NHLF as described previously[17] using a monochromator-based fluorimeter system (OptoFluor; Cairn Research, Kent, UK) and a rapid perfusion system (RSC160, Intracel RSC160, Intracel, Royston, UK) to apply Ca^2+^ (1 or 5 mM), ornithine (5 mM), spermine (1 mM), with or without the calcilytic NPS2143 (20 min pre-incubation, 1 µM). 200 µM ATP was used as a control for cell viability. Background-subtracted 340:380 emission ratios were used to calculate maximal fold-change from baseline.

#### Immunofluorescence staining

NHLF were fixed with 2% paraformaldehyde, permeabilized with 0.1% Triton X-100 and incubated in primary antibodies (mouse CaSR mAb, 1:200, #ab19347; rabbit αSMA polyclonal antibody, 1:100, #ab5694 (both from Abcam); rabbit anti-human anti-Rho associated kinase (ROCK1) polyclonal Ab, 1:500, #21850-1-AP; Proteintech, Manchester, UK) overnight at 4°C after which secondary antibodies were applied (goat anti-rabbit AlexaFluor-488, 1:500, #ab150077; goat anti-mouse AlexaFluor-594, 1:1000, #ab150120; all from Abcam). Cells were mounted using Vectashield with DAPI (Vector laboratories Ltd., Peterborough, UK). Negative controls were performed by omitting primary antibodies and by replacing the CaSR and ROCK1 antibodies with IgG2a isotype control (mouse, 1:100, #02-6200; ThermoFisher, Paisley, UK). Images were captured using Olympus BX61 upright epifluorescence (Olympus UK & Ireland, Essex, UK) or Zeiss LSM880 Airyscan confocal microscope (ROCK1). Cell area and fluorescence intensities were quantified by StrataQuest® (TissueGnostics GmbH, Vienna, Austria) with mean values representing data from 58-1767 regions of interest per condition. Representative images were enhanced to the same brightness and contrast.

#### Fibroblast proliferation, collagen synthesis and IL-8 secretion

BrdU cell proliferation (Abcam, Cambridge, UK), total collagen secretion (Sircol™ soluble collagen assay, collagens type I-V, Biocolor, Belfast, UK) and IL-8 release in NHLF culture supernatant (SimpleStep ELISA kit, Abcam, Cambridge, UK) were quantified according to manufacturer’s instruction. 450 nm (BrdU and IL-8 secretion) or 550 nm (collagen synthesis) absorbance values were measured using a CLARIOstar multi-well plate reader (BMG Labtech Ltd., Aylesbury, UK).

### Mouse studies

#### Assessment of pulmonary remodeling in 15 old mice with targeted CaSR ablation from SM22α^+^ cells

Cre-negative (WT) and ^SM22α^*Casr*^Δflox/Δflox^ (KO) animals were generated by breeding LoxP CaSR mice with SM22α-Cre recombinase mice as described previously.[17] The left lung from 15 month-old animals was formalin-fixed, paraffin-embedded and 5 µm sections were Masson’s trichrome stained according to the manufacturer’s instructions (Polyscience Inc., Northampton, UK). Slides were scanned using an Olympus BX41 microscope (Olympus UK & Ireland, Essex, UK) equipped with QImaging QICAM Fast 1394 color digital camera and an OASIS motorized XYZ stage (Objective imaging Ltd., Cambridge, UK). Histopathological scoring to assess the extent of lung fibrosis was performed using the Ashcroft scale. The amount of collagen deposited was quantified using the software StrataQuest® and expressed as a percentage of lung section surface area (see online supplementary material).

#### Statistics

Metabolomic data were log-transformed with MetaboAnalyst 4.0 using R and Bioconductor packages. Further statistical analyses were carried out as described previously.[21] See online supplementary material.

All other data were analyzed using Prism 7.0 software package (GraphPad Inc.) and are expressed as mean ± SEM. For *in vitro* experiments, data are expressed as a percentage of TGFβ1 treatment. Statistical comparisons were performed using 2-tailed Student’s t-test or ANOVA (with Bonferroni *post hoc* analysis), p-values < 0.05 were considered significant. *N* denotes the number of patients or animals (biological replicates) while *n* represents the number of independent experiments (technical replicates).

## RESULTS

### The CaSR is expressed in the peripheral bronchiolar epithelium and its expression is increased in the neuroepithelial bodies of the human IPF lung

Specific CaSR immunostaining is found in the respiratory epithelium of small peripheral bronchioles, particularly at the apical membranes of ciliated epithelial cells of both control (Figure 1A) and IPF lung tissue (Figure 1, B and Di). CaSR is absent in alveolar regions (Figure S1A). The strongest CaSR expression is seen in the neuroepithelial bodies (NEBs), identified by their known marker, gastrin-releasing peptide (GRP) in serial sections from IPF tissue (Figure 1C). NEBs are more abundant in peripheral bronchioles of IPF patients (Figure 1, B and C), while they are rare in adult control lungs. Above-background CaSR immunoreactivity is found in the interstitium of IPF, but not in control lung (Figure 1, A and B). Immunohistochemistry of serial sections demonstrates almost complete absence of CaSR staining in alpha smooth muscle actin (αSMA)-positive clusters in alveolar (Figure S1A) and bronchiolar regions of IPF lungs (Figure S1B). Confirming the specificity of the signal, no immunoreactivity is observed when the CaSR antibody is pre-absorbed with specific antigenic peptide (Figure 1Dii) or when the primary antibody is replaced with an IgG isotype control (Figure 1Diii) compared with the control tissue (Figure Di).

**Figure 1.**
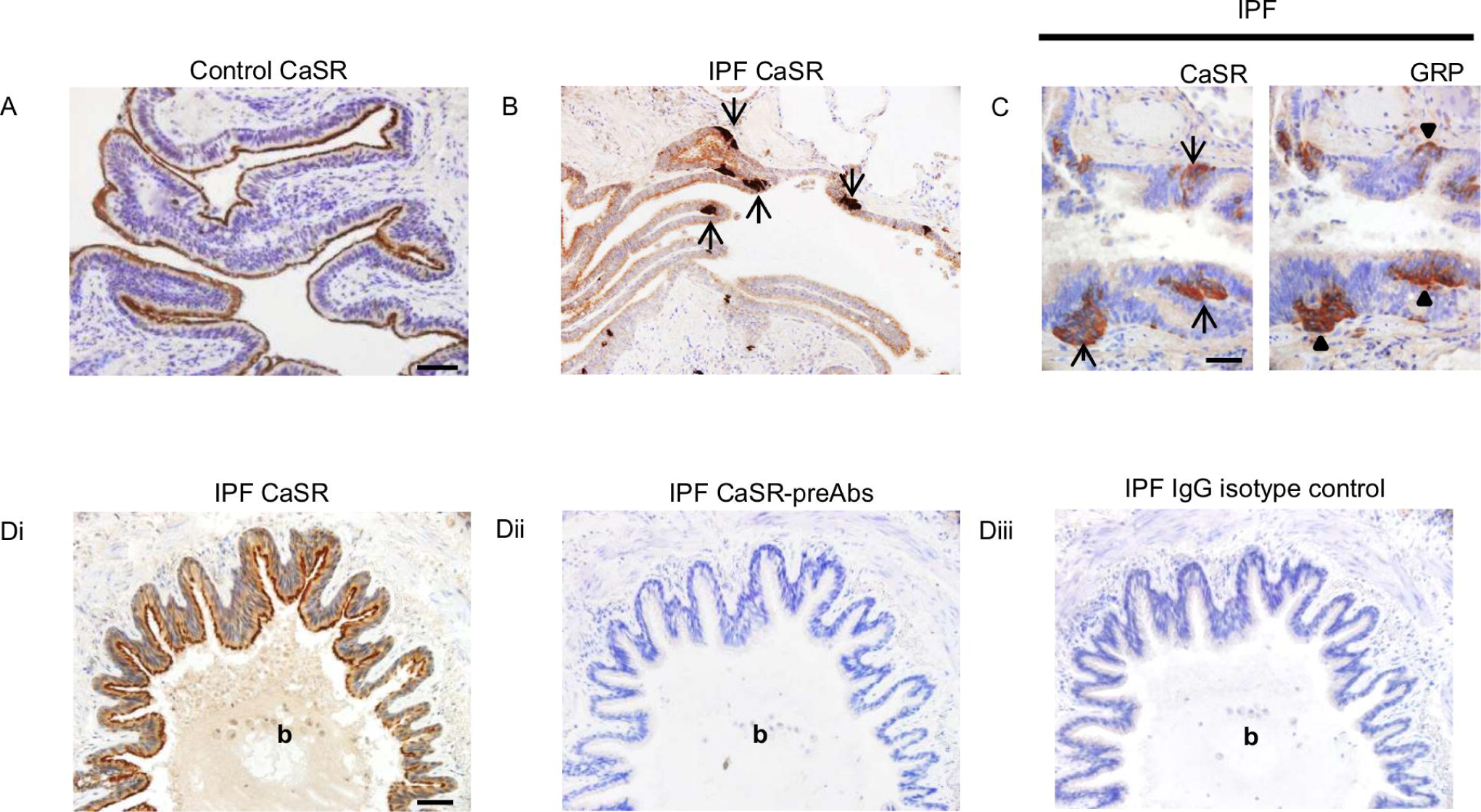
CaSR is expressed in the epithelium of peripheral bronchioles in control and IPF lungs and the number of CaSR-expressing neuroepithelial bodies (NEB) is increased in IPF lungs. **A-D:** immunohistochemistry carried out on paraffin sections of control and IPF lungs. **(A)** CaSR is expressed in the epithelium of peripheral bronchioles in control lungs, **(B)** Overview of a peripheral bronchiole revealing high numbers of grouped NEB-like cells with a strong CaSR immunostaining (*arrows*) in an IPF patient; note that regular bronchiolar epithelial cells also express CaSR. **(Ci, ii)** Serial sections of IPF lung tissue -Grouped CaSR expressing neuroendocrine cells (*arrows*) overlap with cells expressing the human NEB marker gastrin-releasing peptide (GRP; arrowheads). **(Di-iii)** Serial sections of IPF lung tissue - CaSR immunostaining in the airway epithelium **(Di)** is fully abolished after preabsorption of the CaSR antibodies with CaSR antigenic peptide **(Dii)**. No staining can be seen in the epithelium when the primary CaSR antibody is replaced by a non-immune isotype IgG control **(Diii)**. *N* = 5 control donors; 7 IPF donors. *b: lumen of bronchiole* in figures **Di-iii**. Scale bars: 100 µm **(A-B, D)**; 50 µm **(C)**.

### Expression of naturally occurring polyamines is increased in the saliva of PF patients

Principal component analysis (PCA) on saliva samples demonstrates clear clustering of the IPF group, which is distinct from a more diffuse healthy control group (Figure 2A). Assessment of the main sources of variation between the clusters indicates the prominence of the arginine pathway (Figure 2B) and its metabolites (Figure 2C). PF saliva samples exhibit higher levels of arginine (*p*<0.05), ornithine (*p*<0.05) and putrescine (*p*<0.01) compared to controls (Figure 2C). These metabolomic signatures suggest that arginase activity is upregulated in PF saliva.

**Figure 2.**
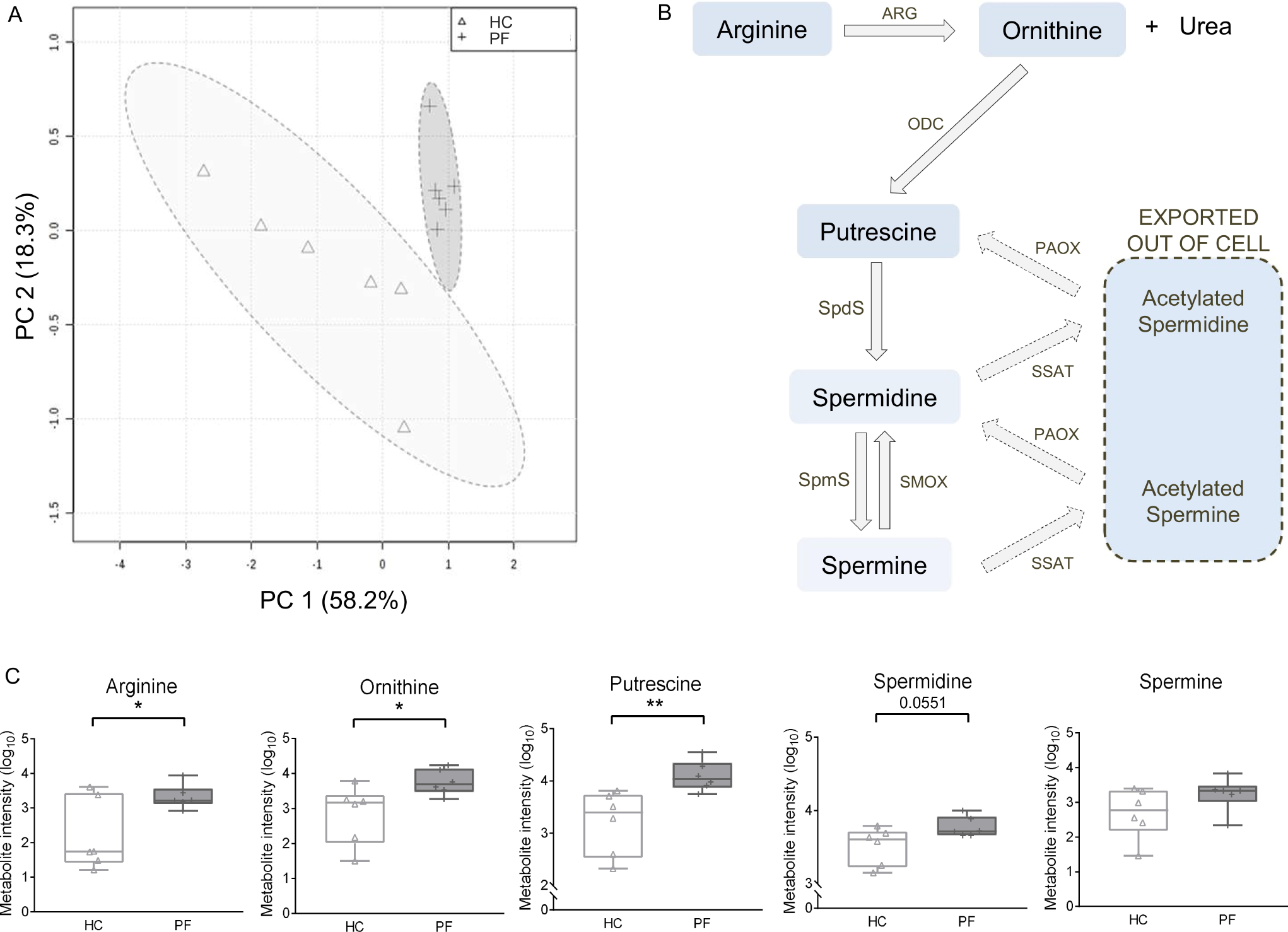
Expression of arginine pathway metabolites is increased in PF patient saliva samples. **(A)** Principal component analysis (PCA) score plot between the main principal components (PC1 and PC2) shows a tight cluster of the PF samples based on their metabolic profile compared to controls with a 95% confidence ellipse drawn for each group (shown by the dotted lines). Assessment of PC1 and PC2, which account for 76.4% of the total variation between the control and PF clusters highlights the prominence of (poly)amines. (**B**) Associated box and whisker plots of metabolites in the arginine-polyamine pathway highlights the differences between healthy and PF profiles. Y-axis: Log_10_ normalization of metabolite intensity. 2-tailed Student’s t test; \**p*<0.05, \*\**p*<0.01. *N* = 6 controls; 6 PF patients. (**C**) Diagram representing the arginine-polyamine metabolic pathway. *HC: control; PF: pulmonary fibrosis; ARG: arginase; ODC: ornithine decarboxylase; SpdS: spermidine synthase; SpmS: spermine synthase; SMOX: Spermine oxidase; POAX: polyamine oxidase; SSAT: spermidine/spermine N1-acetyltransferase 1 (SSAT)*.

### Polyamines upregulated in PF activate the CaSR in human lung fibroblasts

Fibroblasts are the key mediators of the pro-fibrotic response in IPF and crucial to this process is an increase in [Ca^2+^]_i_, which underpins key cellular functions such as gene expression and cell proliferation.[22] Since natural polyamines and basic amino acids are known activators at the CaSR,[12] we tested the hypothesis that polyamines could induce an increase in [Ca^2+^]_i_ in fibroblasts by acting via the CaSR. In NHLF, ornithine and spermine significantly increase [Ca^2+^]_i_ (Figure 3, Ai-Ci and D), as does the CaSR ligand, Ca^2+^. All these effects are completely abolished by calcilytic (*p*<0.001, *p*<0.001 and *p*<0.01, respectively) (Figure 3, Aii-Cii and D) supporting a role for the CaSR in mediating these effects. Immunofluorescence microscopy confirms CaSR expression in NHLF (Figure 3E).

**Figure 3.**
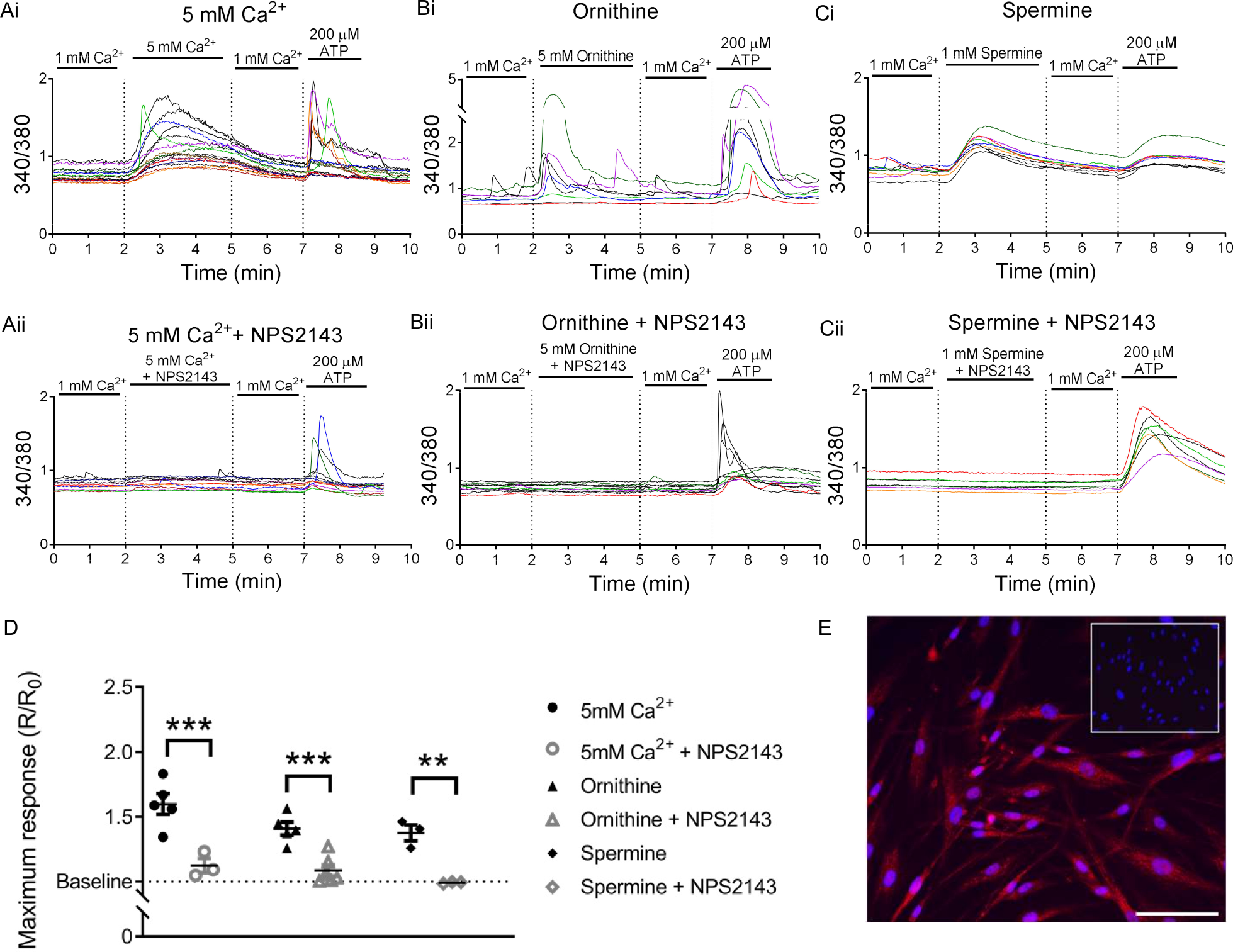
Calcium-sensing receptor (CaSR) is expressed in normal human lung fibroblasts (NHLFs) and is activated by polyamines upregulated in PF. Representative traces (**A-C**) and summary data (**D**) of intracellular calcium levels ([Ca^2+^]_i_) in NHLF in response to: (**Ai**) 5 mM Ca^2+^ _o_ (divalent cation); (**Bi**) 5 mM L-ornithine (basic amino acid); and (**Ci**) 1 mM spermine (polyamine). (**Aii-Cii, D**) Treatment with calcilytic prevents these increases in [Ca^2+^]_i_. Data are shown as mean ± SEM. ANOVA (Bonferroni post hoc test); \*\**p*<0.01, \*\*\**p*<0.001. 5 mM Ca (*n* = 3-6; 108 cells), L-ornithine (*n* = 5-6; 109 cells) and spermine (*n* = 3; 39 cells). (**F**) Representative image of CaSR expression in NHLF (red), nuclei (blue), negative control (insert), *N* = 6 donors. Scale bar: 100 µm.

### TGFβ1 increases CaSR expression and CaSR antagonism reduces TGFβ1 pro-fibrotic responses in human lung fibroblasts

CaSR expression in NHLF is significantly increased by TGFβ1 treatment compared to vehicle-treated cells (Figure 4, A and B, red; Figure S2A). TGFβ1 increases stress fiber formation, ROCK1 and αSMA protein expression; these effects are significantly reduced by the calcilytic (*p*<0.05) (Figure 4, C-F). Calcilytic treatment also significantly decreases TGFβ1-induced proliferation (Figure 4G), soluble collagen (Figure 4H) and IL-8 secretion (Figure 4I) (*p*<0.01, *p*<0.05 and p<0.01, respectively), effects not seen in the absence of TGFβ1 (Figure S2).

**Figure 4.**
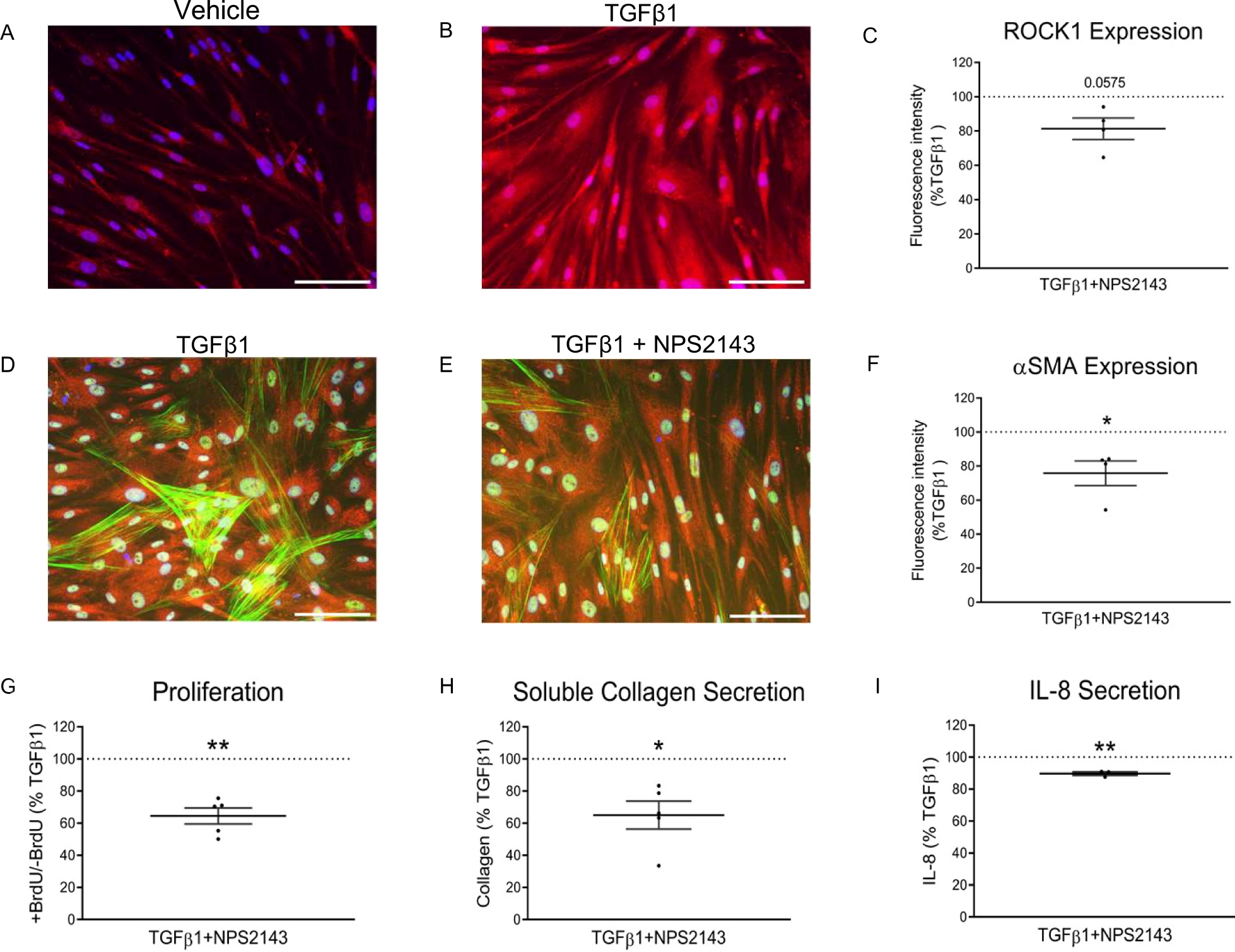
The calcilytic NPS2143 attenuates TGFβ1-induced pro-fibrotic changes in normal human lung fibroblasts (NHLFs). (**A**) NHLFs show low baseline CaSR expression (red). (**B, D**) Treatment with TGFβ1 results in increases CaSR expression and stress fiber formation. (**C**,**F**,**G-I**) The calcilytic NPS2143 inhibits TGFβ1-induced ROCK1 expression (as a surrogate for Rho kinase activation) (**C**), stress-fiber formation (**E**), alpha smooth muscle actin (αSMA) expression (**F**), proliferation (**G**), soluble collagen secretion (**H**), and IL-8 secretion (**I**). Data are normalized to TGFβ1 (which was set at 100%, dotted line) and expressed as mean ± SEM. 2-tailed t-test; \**p*<0.05,\*\**p*<0.01 (**F-I**). *N* = 2-4 donors; *n* = 3-5 independent experiments. Scale bars: 100 µm.

### ^SM22α^*Casr*^Δflox/Δflox^ mice are protected from age-related lung remodeling

The incidence of IPF increases with age, with individuals over the age of 50 being predominantly at risk [7]. Therefore, we investigated whether CaSR signaling could itself lead to increased fibrosis of the ageing lung in the absence of exogenous TGFβ1 administration by assessing the consequences of specific *Casr* gene ablation from SM22α^+^ cells in 15 month old mice as a model of naturally occurring, age-related fibrosis. This age of the mice correlates with approximately 50-60 years in humans [23]. Masson’s trichrome stained lung sections from 15 month-old WT mice reveal significantly more extensive fibrosis, thicker interstitial walls and damage to the lung architecture when compared with age-matched KO mouse lung sections (*p*<0.05) (Figure 5, A-C). Absence of CaSR from SM22α^+^ cells is also associated with an overall reduction in collagen deposition around the small airways with the greatest reduction observed 10 µm from the airway lumen (*p*<0.05) (Figure 5D).

**Figure 5.**
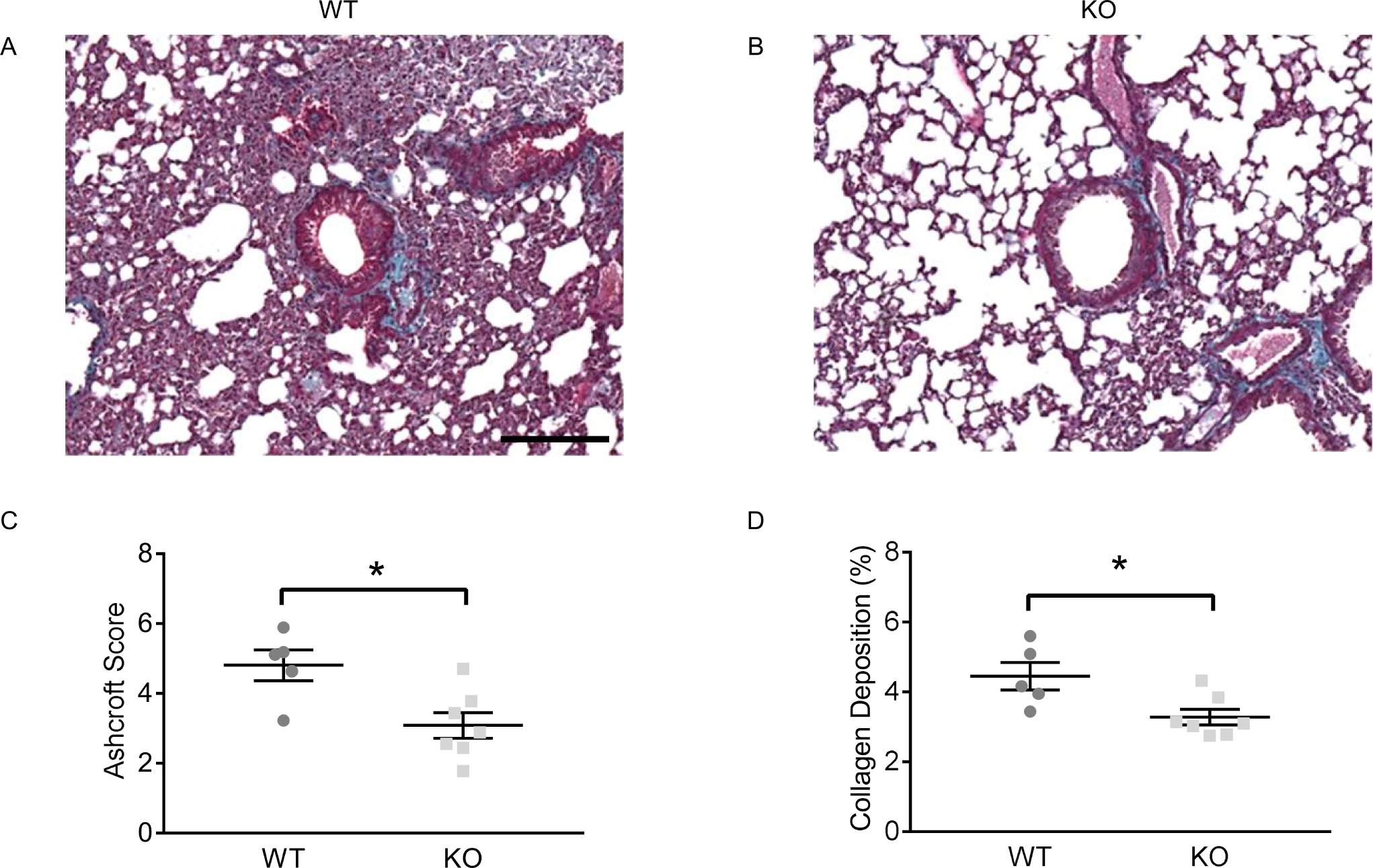
Targeted CaSR deletion from SM22α^+^ cells protects mice from age-related lung fibrosis. (**A-B**) Histology of lungs from 15 month-old WT mice shows significant fibrotic remodeling compared to lungs from age-matched mice with targeted CaSR deletion (KO); quantified using the Ashcroft score (**C**). (**D**) Collagen deposition is significantly decreased around the airway lumen of KO mice compared to WT. CaSR gene was selectively deleted downstream of the SM22α promoter using the Cre-Lox system; Cre-negative (WT) SM22^α^, *Casr* ^Δflox/Δα^ (KO). Collagen deposition was quantified using StrataQuest - collagen expression (density of blue pixels) is presented as a percentage of the total lung surface area up to 10 µm away from airway lumen (%). Data are expressed as mean ± SEM. 2-tailed Student’s t test, * *p*<0.05. *N* = 5 (WT); 7 (KO). Scale bar: 100 μm (**A-B**). *SM22*α: *smooth muscle-associated protein of 22kDa*.

## DISCUSSION

Recent evidence demonstrates the importance of GPCR signaling in IPF. This study highlights a role for the GPCR CaSR in IPF etiology, and the possibility to use CaSR blockers, calcilytics, as a novel treatment for IPF.

We found the most abundant CaSR expression in the airway epithelium and in NEBs. This expression pattern is consistent with the receptor integrating signals derived from environmental cues, endogenous compounds, oxidative stress levels and energy status.[24] CaSR activation mediates wound-healing in human airway epithelial cells by increasing proliferation and migration *via* [Ca^2+^]_i_ mobilization, activation of extracellular signal-regulated kinase (ERK1/2) and Rho kinase signaling,[15] thereby resulting in the activation of pro-fibrotic fibroblast regulators.[9] We hypothesize that inappropriately sustained exposure to environmental pollutants and biological agents such as smoke, urban particulate matter and viruses, all of which contain a plethora of polycations/polyamines acting at the CaSR,[16] provides the stimulus for both initiation and progression of the fibrotic cascade *in vivo*. Therefore, aberrant CaSR activation in epithelial cells could account for some of the histopathological features of IPF, namely airway epithelial cell hyperplasia within honeycomb cysts, and bronchiolization of distal airways and alveolar spaces.[25]

Although many consider alveolar epithelial cells as the initial site of injury in IPF, our study reports an increase in the number of NEBs in the lung of IPF patients. NEBs serve diverse functions during fetal, perinatal, and postnatal life,[26] including development/growth of surrounding the airway epithelium,[26] and sensing/transducing hypoxic,[27] mechanical,[24] chemical,[28] and likely other stimuli from the environment. They also represent a unique stem cell niche involved in airway epithelium repair.[29] Particularly relevant to our findings is the strong surface membrane expression of fully functional CaSR reported in mouse NEBs.[28] Among several other bioactive substances, human NEBs synthesize, store and secrete GRP in health and disease,[30] which is implicated in (myo)fibroblast proliferation, collagen synthesis and alveolar wall thickening in pulmonary fibrosis.[10, 30] We speculate that the sensory ability and neuropeptide release from highly proliferating NEB cells may be amplified in IPF due to significant changes in ECM stiffness and oxygen availability. These changes may be further potentiated by CaSR activation with attendant increase in [Ca^2+^]_i_ and GRP release, exacerbating the fibrotic process.

IPF diagnosis is hampered by the absence of non-invasive disease biomarkers. Our findings show that PF patient saliva samples exhibit an upregulation of the arginine pathway metabolites compared to healthy controls. These results are consistent with previous studies showing increased arginase expression and activity in human and rodent models of PF[31] and with increased ornithine-derived metabolites, putrescine and spermidine, in IPF lung tissue.[14] Moreover, in the arginine metabolic cycle, ornithine is the rate-limiting step in polyamine/polypeptide synthesis[32] and is an activator of the CaSR, as are the ornithine-derived polyamines, putrescine, spermidine and spermine. Since salivary biomarkers correlate with serum biomarkers in several systemic diseases,[33] our results suggest that saliva is a credible biofluid for disease detection and for patient stratification.

One of the sources of the myofibroblast population found within IPF fibrotic foci are activated resident fibroblasts, which proliferate and differentiate into matrix secreting, αSMA^+^, myofibroblasts. Although interstitial CaSR is relatively low/absent in lung tissue, here we show that TGFβ1 increases CaSR expression in NHLF. Several TGFβ1-mediated pathways are implicated in αSMA stress fiber formation including ERK, Akt and Rho-kinase activation.[11] CaSR activation in NHLF induces [Ca^2+^]_i_ mobilization and cytoskeletal rearrangements, which drive pro-fibrotic processes. Calcilytics reduce these effects, in addition to potentially increasing the intracellular cAMP pool,[16] thereby acting as novel anti-fibrotic agents. In addition, TGFβ1 induced increases CaSR expression which is likely to create a positive feedback loop driving disease progression. Calcilytics have the potential to break this cycle and attenuate key pathological hallmarks of IPF: alteration in cell morphology, fibroblast proliferation, myofibroblast accumulation, ECM deposition and inflammation.

Although inflammation is thought to play a limited role in established IPF, it may be important in the early stages of the disease.[34] IL-8 is a key mediator of IPF-related processes such as cell fibrogenicity;[35] as little as 1 pg/ml increase in serum IL-8 correlates with increased risk of death by 6.7%.[36] Furthermore, IL-8 is a potential IPF biomarker; its levels are increased in the bronchoalveolar fluid, serum and sputum samples of IPF patients.[37] The current study shows that calcilytic treatment significantly reduces IL-8 secretion, suggesting that IL-8 measurements could be used to assess calcilytic efficacy in the clinic.

IPF is seen an accelerated form of ageing,[38] and patient survival is severely reduced in older individuals.[7] Our results show that selective Casr ablation from SM22α-positive cells protects mice from age-induced fibrosis and preserves normal lung architecture. These results suggest that the CaSR also plays a role in age-related pulmonary fibrosis and corroborates the idea that receptor antagonism might be beneficial in the treatment of IPF in elderly patients.

Existing IPF treatments are not curative and are associated with severe side effects. Systemic calcilytics were initially developed for treating osteoporosis; although safe and well-tolerated in clinical trials, their development for osteoporosis was discontinued because they failed to improve bone mineral density and caused mild, asymptomatic hypercalcemia.[39] Topical delivery of calcilytics has being tested in surrogate models of inflammatory lung disease without any systemic effects,[16, 19] suggesting that they could be developed as an addition to the existing IPF therapeutic armamentarium.

IPF patients often suffer from several coexisting lung conditions such as emphysema and PAH that are not targeted by either pirfenidone or nintedanib.[5] Previous studies show that calcilytics prevent profibrotic changes and remodeling in preclinical models of PAH and COPD[18, 19] suggesting the possibility of using a single agent for treating both IPF and its comorbidities, further improving patient outcomes. Our data show that the CaSR is expressed in many cells of the IPF lung. By targeting the CaSR and its associated intracellular pathways in multiple cell types, calcilytics have the potential to act upstream of the fibrotic process (Figure 6) and should halt disease progression, thereby conferring a unique mechanistic advantage over existing approved medications.

**Figure 6.**
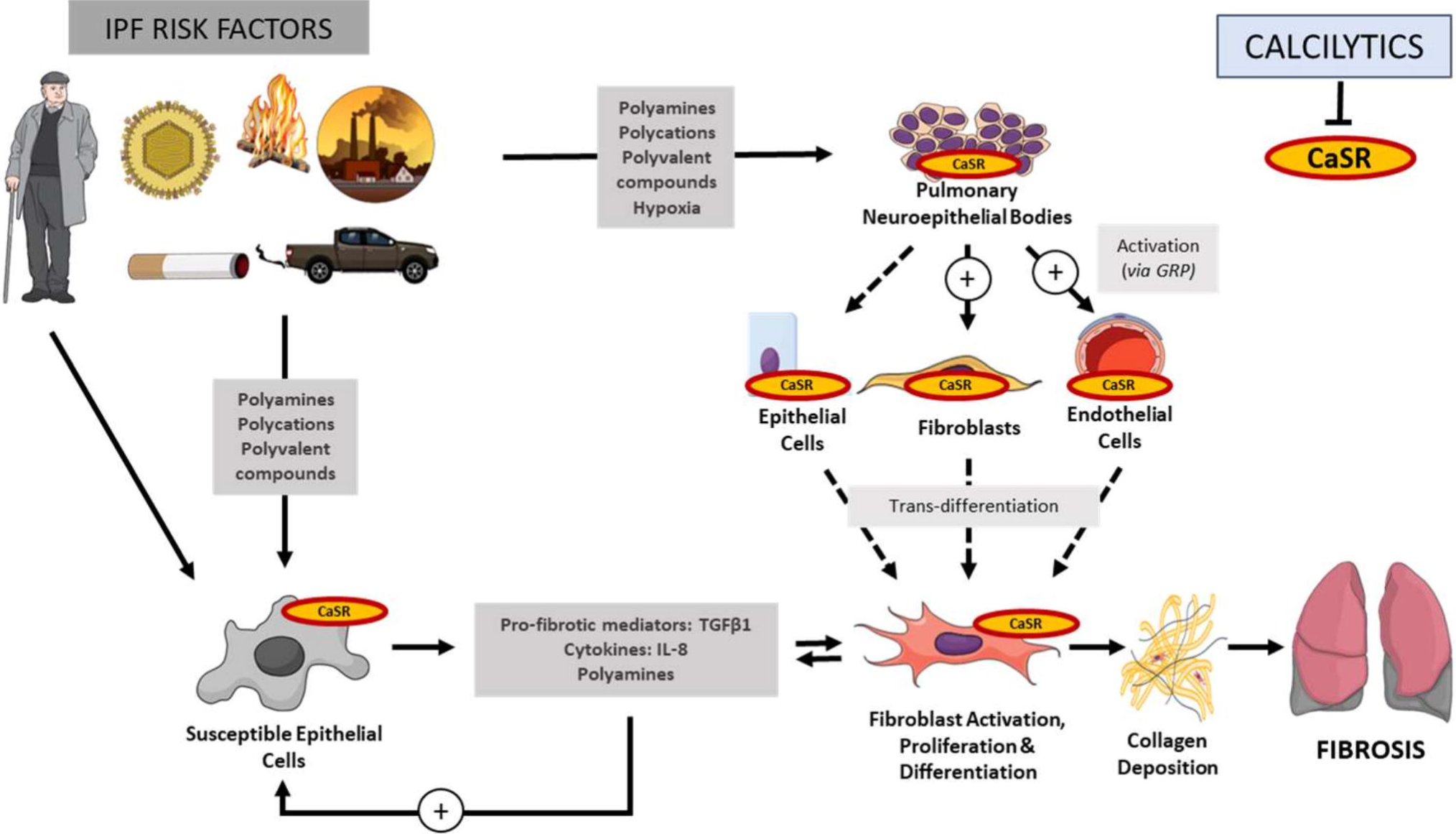
Schematic diagram illustrating the role of the CaSR in the pathogenesis of IPF. Environmental triggers (e.g. urban particulate matter, cigarette smoke, viral particles) contain polycations which activate the CaSR, as do the polyamines associated with the arginase pathway. Polycations and polyamines activate the CaSR in the bronchiolar epithelium and neuroepithelial bodies (NEBs), leading to release of pro-fibrotic factors. The most researched (I)PF-related cytokine, TGFβ1, drives fibroblast activation, proliferation and differentiation into myofibroblasts resulting in excessive ECM deposition. Furthermore, fibroblasts also secrete cytokines which facilitate a positive feedback loop resulting in epithelial cell activation thereby exacerbating the fibrotic process. NEBs act as a potential stem-cell niche for epithelial cells and release profibrotic neuropeptides e.g. gastrin-releasing peptide (GRP) which activate fibroblasts and endothelial cells. These CaSR-expressing cells are also cellular mediators of common IPF comorbidities, pulmonary arterial hypertension and chronic obstructive pulmonary disease. Senescent cell accumulation and increased TGFβ1 expression are key hallmarks of ageing that increase susceptibility of epithelial cells to injury thereby reinforcing the fibrotic pathway.

## Data Availability

All data relevant to the study are included in the article or uploaded as supplementary information

## ACKNOWLEDGEMENT

We wish to thank Dr J. Dally and Dr R. Moseley for their technical advice, and Dr Wenhan Chang for the LoxP CaSR mice.

## CONTRIBUTORS

KLW and DR designed the study. KLW, BM, RTB, LV, RPDA, PH conducted the experiments. MS, SCB generated the WT and KO mouse line. KLW, RTB, LAJM, DA analyzed the data. Patient samples were provided by RA, BH-G, DA, LAJM, KEL. KLW, DR, DA wrote the manuscript, CJC, KEL, LAJM, BH-G, LV, PJK, CJC, JPTW, MS, SCB, provided feedback on drafts. All authors read and approved final draft of the manuscript.

## PATIENT CONSENT FOR PUBLICATION

Obtained

## ETHICAL APPROVAL

### Human tissue collection

Human biopsy samples were taken from IPF patients for diagnostic purposes and obtained with the approval of Cardiff University Ethics Committee and Cardiff and Vale UHB Research and Development department. The control lung sections originated from tissue identified as ‘normal’ by a thoracic pathologist, after lung tumor resections. Samples obtained in compliance with the UK human tissue act 2004.

### Patient saliva sample collection

The study “Novel Technologies for Diagnosing and Monitoring Respiratory Diseases” obtained regional ethical approval from Hywel Dda University Health Board (16/WA/0036). The study was conducted in accordance with the Helsinki principles. Data were link-anonymized before analysis. Eligibility included confirmed diagnosis of PF at the time of sample collection. Control samples were collected from spouses accompanying patients attending clinics.

### Mouse experiments

All animal procedures are in line with the regulations of the Animals (Scientific Procedures) Act 1986, monitored by the Home Office (UK) and Cardiff University Ethical Committee, and conform to the ARRIVE guidelines.

## PROVENANCE AND PEER REVIEW

Not commissioned, externally peer reviewed.

## DATA AVAILABILITY STATEMENT

All data relevant to the study are included in the article or uploaded as supplementary information.

